# Food Insecurity amid COVID-19 Lockdowns: Assessing Sociodemographic Indicators of Vulnerability in Harar and Kersa, Ethiopia

**DOI:** 10.1101/2023.01.31.23284545

**Authors:** Jonathan A. Muir, Merga Dheresa, Zachary J. Madewell, Tamirat Getachew, Gezahegn Mengesha, Cynthia G. Whitney, Nega Assefa, Solveig A. Cunningham

## Abstract

**Objective:** The COVID-19 pandemic was associated with widespread social disruptions, as governments implemented lockdowns to quell disease spread. To advance knowledge of consequences for households in lower-income countries, we examine food insecurity during the pandemic period.

**Design:** Cross-sectional study using logistic regression to examine factors associated with food insecurity. Data were collected between August and September of 2021 through a Health and Demographic Surveillance System (HDSS) using a survey instrument focused on knowledge regarding the spread of COVID-19; food availability; COVID-19 related shocks/coping; under-five child healthcare services; and healthcare services for pregnant women.

**Setting:** The study is set in two communities in Eastern Ethiopia, one rural and one urban.

**Participants:** A random sample of 880 households residing in Kersa and Harar.

**Results:** Roughly 16% of households reported not having enough food to eat during the pandemic, an increase of 6% since before the pandemic. After adjusting for other variables, households were more likely to report food insecurity if they were living in an urban area, were a larger household, had a family member lose employment, reported an increase in food prices, or were food insecure before the pandemic. Households were less likely to report food insecurity if they were wealthier or had higher household income. **Discussion** After taking other characteristics into consideration, households in urban areas were at higher risk for food insecurity. These findings point to the need for expanding food assistance programs to more urban areas to help mitigate the impact of lockdowns on more vulnerable households.

## INTRODUCTION

COVID-19 was associated with one of the largest disruptions to life in the 21^st^ century. Beyond immediate health effects,[1] the pandemic also had social and economic implications as government-enforced lockdowns were implemented with the goal of mitigating the pace and severity of the disease.[2] To comply with these lockdowns, many businesses had to close in-person offices or shops; for some, this meant closing permanently. Healthcare providers also had to restrict in-person access to patients and/or decrease their services. These lockdowns are also thought to have affected the health of children by limiting families’ ability to access food—especially in isolated regions of resource-limited countries.[3–13] Information on the extent to which households living in non-western countries have experienced resource restrictions and other hardships remains limited due to incomplete or nonexistent population health surveillance.[14] We utilized data from an existing Health and Demographic Surveillance System (HDSS) with catchment areas situated in an urban area of the Harari People’s National Regional State and a rural area within the district of Kersa in Eastern Ethiopia. We employed a social vulnerabilities framework[15, 16] that considered both social inequalities and place inequalities as components of vulnerability to analyze food insecurity in the context of the pandemic.

## BACKGROUND

Food security is the condition in which “all people, at all times, have physical, social, and economic access to sufficient, safe, and nutritious food that meets their dietary needs and food preferences for an active and healthy life.”[17] It has been described across four dimensions: food availability, food accessibility, food utilization and food stability.[6] It is anticipated that households around the world, especially in resource-limited countries, have experienced increases in food *insecurity* due to the social and economic repercussions of COVID-19.[3–13] Pathways through which lockdowns may exacerbate food insecurities include movement restrictions, food supply chain disruptions, disruptions to informal markets, food price increases, reductions in individual or household income due to job loss or business closure, and loss of access to government food programs.[7] Although mortality directly related to COVID-19 is lower among children and pregnant women compared to older adults, reductions in access to basic nutrition, coupled with disruptions in access to preventative maternal and child healthcare, may increase mortality and morbidity among young children and pregnant women.[4, 18–22] Particular concern has been raised regarding health consequences of pandemic-related lockdowns in sub-Saharan Africa.[8]

### COVID-19 Lockdowns in Sub-Saharan Africa and Ethiopia

In sub-Saharan Africa, national governments have taken substantial efforts to quell the spread of COVID-19.[2, 23] Implementation of physical distancing, sanitary measures, and encouragement of personal responsibility have been used to help slow the spread of disease. In particular, communities have been tasked with monitoring self-isolation measures, physical distancing, and quarantining. A notable challenge that many sub-Saharan Africa countries have likely faced during pandemic-related lockdowns is food insecurity.[3–9] Many countries in Sub-Saharan Africa rely on food imports, particularly in urban areas; lockdowns and related border closures may compromise these food supply chains, leading to food shortages and price increases.[8] Coupled with on-going climate change, pests, and emerging crop diseases, it is anticipated that there will be many who are indirectly hurt by the pandemic in addition to those directly affected by COVID-19. This is likely the case Ethiopia, where the government responded with stringent measures to mitigate the pace and severity of COVID-19.[24]

Mitigation efforts in Ethiopia included establishing lockdowns, enforcing social distancing, and placing emphasis on hygiene protocols; these efforts were initially adopted on March 16^th^ of 2020, tightened on March 20^th^, and a five-month state of emergency was declared on April 10^th^.[24] Yet, social and economic disparities across different sociodemographic groups and geospatial inequalities may have resulted in uneven implementation of these efforts.[11] Populations in urban centers may have been better equipped to receive information about these directives compared to rural populations that face communication barriers.[25] Rural populations likely confronted challenges with implementing some measures due to limited access to handwashing facilities or lack spare rooms in their dwellings for social distancing or quarantine; however lower population density may have aided in social distancing between households.[25] Social and economic disparities may also have resulted in differential vulnerability to adverse consequences of mitigation efforts.

### Vulnerability to Food Insecurity in Ethiopia

Vulnerability, broadly defined, is the cumulative effect of cultural, economic, institutional, political, and social processes that modify socioeconomic differences in the experience of and recovery from hazards.[15, 26] In the context of disasters, it is often not the hazard itself that creates the disaster; rather, the disaster is the impact on individual and community coping patterns and the inputs and outputs of social systems.[27–29] Social vulnerability is partially the result of social disparities that shape or influence the susceptibility of different groups to hazards while also controlling their capacity to respond.[15, 16] Factors often associated with vulnerability include demographic characteristics such as age, ethnicity, race, and sex; socioeconomic status (e.g., income, wealth, employment, and/or education); household composition (e.g., presence of children or elderly); and housing and transportation.[30] However, social vulnerability also involves place disparities stemming from characteristics of communities and the built environment that contribute to the vulnerability of place. For example, differential availability of scarce resources between urban and rural areas may exacerbate social vulnerabilities to hazards.[16, 31, 32] To understand the broader consequences of the pandemic, it is important to consider economic, political, and social markers of vulnerability at both the individual and community level.[23]

In Ethiopia, food insecurity was prevalent prior to the onset of the pandemic with those already burdened by poverty at greater risk for experiencing food insecurity; it is anticipated that COVID-19 has exacerbated this vulnerability.[7, 11] Households hit with an economic shock such as a job loss or loss of income during the pandemic are likely at high risk for experiencing food insecurity, as they must reduce expenditures to adapt to the economic shock.[9, 33] Disadvantaged groups such as women and children, people working in the informal sector or unemployed, those with limited financial resources, those with medical conditions, and those relying on emergency food aid are also at high risk.[23, 34] Rural populations in Ethiopia are at elevated risk for food insecurity as climate change, severe drought, conflict, and environmental degradation have culminated in societal shocks affecting livelihoods, particularly for farmers.[16, 35, 36] However, understanding geospatial differences in vulnerability is complicated because international and national food programs were established to provide food aid to needy populations, especially in rural areas, prior to the onset of the pandemic.[37] Given their strong emphasis on addressing food insecurity in rural areas, these programs may not reach food insecure households living in urban areas.[37, 38] Differential vulnerability requires targeted, context specific interventions to address ancillary consequences of the pandemic such as increases in food insecurity.[39] To help inform such interventions and increase their precision and efficacy, we analyze demographic, economic, and social factors associated with food insecurity during the pandemic.

## METHODS

### Study setting

The setting for this study is a predominantly rural area in the district of Kersa and an urban area in the Harari People’s National Regional State in Eastern Ethiopia.[40, 41] The rural area consists of 24 kebeles (a neighborhood or ward representing the lowest level of local government), and covers 353 km^2^ with a population of 135,754 in 25,653 households. The urban area consists of 12 kebeles, a population of 55,773 in 14,768 households, across 25.4 km^2^.[41] The population has been followed through a Health and Demographic Surveillance System since 2012, with demographic and health-related information regularly collected (see Figure 1).

**Figure 1.**
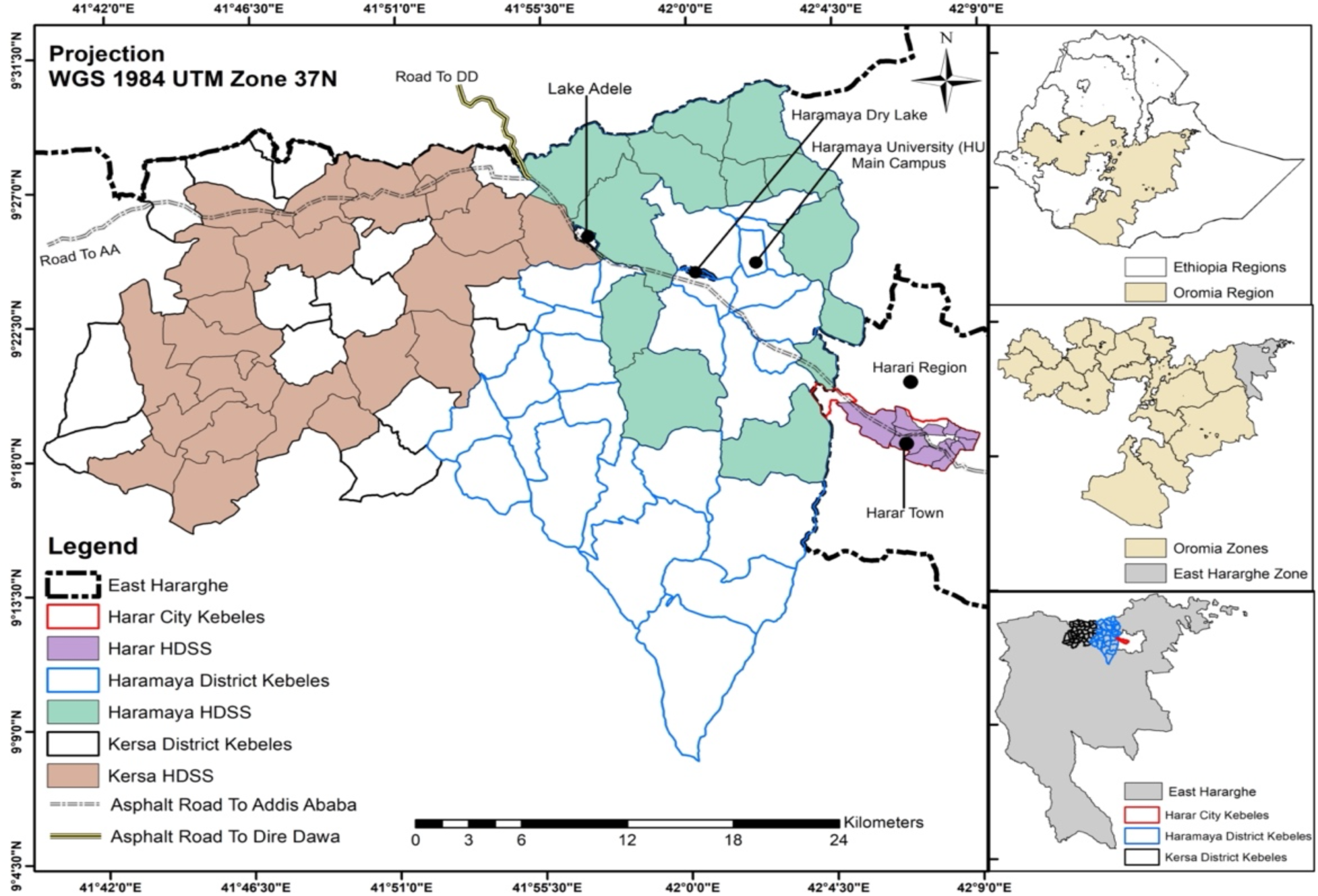
The Harar and Kersa Health and Demographic Surveillance Systems (HDSS) within East Hararghe, Oromia, Ethiopia. The smaller map panels on the right identify the location of the HDSS catchment areas within the East Haraghe Zone of the Oromia region in Ethiopia. The HDSS catchment in Haramaya (depicted in green) was in development during the data collection period, so households from this catchment were not included in this study.

### Study design

This study is part of a broader investigation within the Child Health and Mortality Prevention Surveillance (CHAMPS) network to understand the implications of COVID-19 lockdowns for child health and mortality[33, 42]. We developed and administered a short quantitative instrument designed to understand how the pandemic and related lockdowns may have affected livelihood, food availability, and healthcare. Households were selected using simple random sampling. The sample size was set to 440 each from the rural (Kersa) and urban (Harar) catchment areas (total sample size of 880 households). The sample size was specified to detect prevalence of changes in accessing care. *A priori* specifications were 50% of the population experiencing changes, 95% CI, precision of 0.05 and non-response adjustment of 10%.

The data collection instrument was organized into five sections: knowledge regarding the spread of COVID-19; food availability; COVID-19 related shocks/coping; under-five child healthcare services; and healthcare services for pregnant women (see Supplementary Methods) [43]. Questions in the survey instrument related to hardships associated with the pandemic period asked respondents to consider whether they had experienced hardships since March of 2020. Data were collected by HDSS enumerators in August and September 2021 through tablet-based in-person interviews with heads of households or their spouse. Households’ responses were linked with their data from the most recent HDSS round, which had occurred in 2021, to incorporate information on age, sex, occupation, and education of the head of household; the number of children under 5 years of age and the number of elderly adults (over 60 years) in the household; and household assets and residence construction materials. All data were collected digitally using tablets that were programmed with the corresponding survey instruments.

Data quality assurance and cleaning followed standard operating procedures for the HDSS[40, 44]. Inconsistent or missing data were flagged for data collectors to correct. Field supervisors and the field coordinator selected a random sample of questionnaires for re-visits to verify the recorded information. Implementation of the module was approved by the Institutional Health Research Ethics Review Committee (IHRERC); approval reference number Ref.No.IHRERC/127/2021.

### Measures

The variable of interest is *food insecurity*, indicating that a household reported not having enough food to eat at any point since March 2020 based on responses to the question: *“Since the beginning of the COVID lockdown, has it happened that your household did not have enough food to eat?”* The variable is coded as 0 = no, 1 = yes. Additional variables identified the reasons that a household was unable to get enough food, with respondents able to report multiple reasons: *couldn’t afford to buy more food; couldn’t go out to buy food, afraid to go out to buy food; couldn’t get groceries or meals delivered;* and *stores did not have the food I wanted*.

Right-hand side variables were information on the head of household and characteristics of the household and its homestead. Head of household variables were *age* (coded as 1 = less than 40 years, 2 = 41 to 44 years, 3 = 45 to 60 years, and 4 = sixty years and above); *sex* (coded as male = 0 and female = 1); *ethnicity* (coded as Amhara = 1 (reference) and other = 2, which included Oromo, Somali, Gurage, Harari and Tigray); *education* (coded as no formal education = 0 (reference), any level of education = 1); *occupation* (coded as 1 = farmer (reference), 2 = student, 3 = professional, 4 = sales, 5 = housewife, 6 = daily laborer, 7 = other employment, 8 = unemployed/retired). Household variables were *urbanicity* (coded as urban (Harar) = 1 and rural (Kersa) = 0); *household size* (coded as 1 = 1-2 individuals (reference), 2 = 3-4 individuals, 3 = 5-6 individuals, 4 = 7-8 individuals, 5 = 9+ individuals); *children under 5* (coded as 0 = no, 1 = yes); *adults over age 60* (coded as 0 = no, 1 = yes); monthly *income* (coded into quartiles as 1 = 0-1,200 Birr Ethiopia Birr (reference), 2 = 1,201-2,000 Birr, 3 = 2,001-3,000 Birr, 4 = 3,001-4,600 Birr, and 5 = 4,600+)^1^; *wealth index* based on a list of household assets (coded as an 1 = poorest (reference), 2 = poorer, 3 = middle, 4 = richer, 5 = richest)^2^; *job loss* by any household member since March of 2020 (coded as 0 = no, 1 = yes); *food price increase* noticed by the household after March of 2020 (coded as 0 = no, 1 = yes); and *pre-COVID food insecurity* (coded as 0 = no, 1 = yes)^3^.

### Analytic Strategy

Data cleaning and analyses were performed using R version 4.2.0.[45] Education and occupation related data were missing for nine household heads and observations for these households were removed from analyses that included these variables. Data on pre-pandemic food insecurity was missing for one household. There were no other missing values for the variables used.

Descriptive statistics were generated to assess the prevalence of food insecurity and other hardships and sociodemographic characteristics. Pearson’s Chi-square tests were used for preliminary evaluation of associations between household characteristics and food insecurity. Unadjusted logistic regression models were used to analyze associations with food insecurity (results available upon request). Adjusted logistic regression was then used to control for other characteristics.

Several pathway-specific models were examined prior to fitting the final model and are available upon request. The variables described above were selected for inclusion in the final adjusted logistic regression because there was theoretical justification for association with food insecurity. We assessed model fit with comparison of AIC and BIC scores—both scores were the smallest for the fully adjusted model, indicating that this model had superior fit compared to simpler or pathway-specific models. Results are reported as Adjusted Odds Ratios (AOR) with 95% confidence intervals (95% CI) and visualized with forest plots.[46, 47] Given anticipated differences in demographic, economic, and social characteristics between households living in urban vs. rural areas, effect modification was evaluated using interaction terms; however, the results of these analyses were not statistically significant so they were not retained. Finally, a secondary analysis was conducted wherein only those respondents who reported that their household had no difficulty obtaining sufficient food prior to March of 2020 were included. This analysis examined factors associated with the change from food secure prior to COVID-19 to food insecure during the pandemic. For this analysis, some variables from the primary analysis such as ethnicity could not be included due to lack of variation; other variables required recoding to due to sample size limitations.

## RESULTS

A majority of households (63.3%) were headed by a male household member (Table 1). Average age of the household head was 43 years with a standard deviation of 15. Oromo and Amhara were the most common ethnicities, with 551 (62.6%) household heads identifying as Oromo and 222 (25.2%) as Amhara. A majority (57.9%) of household heads had obtained at least some education; their most common occupations were farmer (36.4%) and professional (18.3%). The majority of households resided in urban areas (62.8%). The median number of household members was 5 with a standard deviation of 2.4. Roughly a third of households reported having at least one child under the age of 5. About a quarter of households had at least one adult family member over the age of 60. About 10% of households reported that they were unable to obtain sufficient food prior to the onset of the pandemic; a majority of these households (94%) continued to report food insecurity during the pandemic.

**Table 1:**
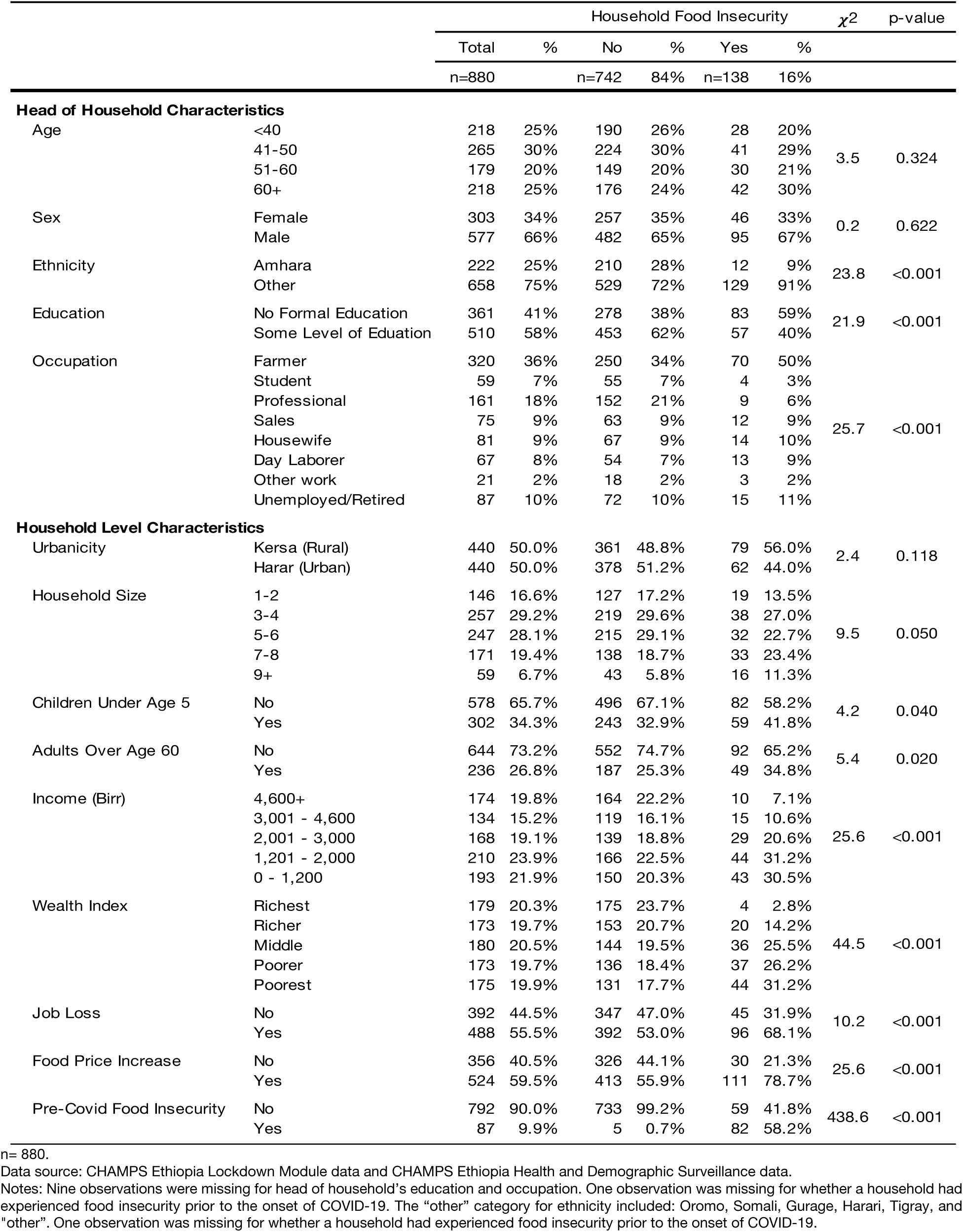
Descriptive Statistics and Chi-Square Values for HDSS Households

| | | Household Food Insecurity | | | | | | $\chi^2$ | p-value |
| --- | --- | --- | --- | --- | --- | --- | --- | --- | --- |
|  |  | Total | % | No | % | Yes | % |  |  |
|  |  | n=880 |  | n=742 | 84% | n=138 | 16% |  |  |
| Head of Household Characteristics |  |  |  |  |  |  |  |  |  |
| Age | <40 | 218 | 25% | 190 | 26% | 28 | 20% | 3.5 | 0.324 |
|  | 41-50 | 265 | 30% | 224 | 30% | 41 | 29% |  |  |
|  | 51-60 | 179 | 20% | 149 | 20% | 30 | 21% |  |  |
|  | 60+ | 218 | 25% | 176 | 24% | 42 | 30% |  |  |
| Sex | Female | 303 | 34% | 257 | 35% | 46 | 33% | 0.2 | 0.622 |
|  | Male | 577 | 66% | 482 | 65% | 95 | 67% |  |  |
| Ethnicity | Amhara | 222 | 25% | 210 | 28% | 12 | 9% | 23.8 | <0.001 |
|  | Other | 658 | 75% | 529 | 72% | 129 | 91% |  |  |
| Education | No Formal Education | 361 | 41% | 278 | 38% | 83 | 59% | 21.9 | <0.001 |
|  | Some Level of Education | 510 | 58% | 453 | 62% | 57 | 40% |  |  |
| Occupation | Farmer | 320 | 36% | 250 | 34% | 70 | 50% | 25.7 | <0.001 |
|  | Student | 59 | 7% | 55 | 7% | 4 | 3% |  |  |
|  | Professional | 161 | 18% | 152 | 21% | 9 | 6% |  |  |
|  | Sales | 75 | 9% | 63 | 9% | 12 | 9% |  |  |
|  | Housewife | 81 | 9% | 67 | 9% | 14 | 10% |  |  |
|  | Day Laborer | 67 | 8% | 54 | 7% | 13 | 9% |  |  |
|  | Other work | 21 | 2% | 18 | 2% | 3 | 2% |  |  |
|  | Unemployed/Retired | 87 | 10% | 72 | 10% | 15 | 11% |  |  |
| Household Level Characteristics |  |  |  |  |  |  |  |  |  |
| Urbanicity | Kersa (Rural) | 440 | 50.0% | 361 | 48.8% | 79 | 56.0% | 2.4 | 0.118 |
|  | Harar (Urban) | 440 | 50.0% | 378 | 51.2% | 62 | 44.0% |  |  |
| Household Size | 1-2 | 146 | 16.6% | 127 | 17.2% | 19 | 13.5% | 9.5 | 0.050 |
|  | 3-4 | 257 | 29.2% | 219 | 29.6% | 38 | 27.0% |  |  |
|  | 5-6 | 247 | 28.1% | 215 | 29.1% | 32 | 22.7% |  |  |
|  | 7-8 | 171 | 19.4% | 138 | 18.7% | 33 | 23.4% |  |  |
|  | 9+ | 59 | 6.7% | 43 | 5.8% | 16 | 11.3% |  |  |
| Children Under Age 5 | No | 578 | 65.7% | 496 | 67.1% | 82 | 58.2% | 4.2 | 0.040 |
|  | Yes | 302 | 34.3% | 243 | 32.9% | 59 | 41.8% |  |  |
| Adults Over Age 60 | No | 644 | 73.2% | 552 | 74.7% | 92 | 65.2% | 5.4 | 0.020 |
|  | Yes | 236 | 26.8% | 187 | 25.3% | 49 | 34.8% |  |  |
| Income (Birr) | 4,600+ | 174 | 19.8% | 164 | 22.2% | 10 | 7.1% | 25.6 | <0.001 |
|  | 3,001 - 4,600 | 134 | 15.2% | 119 | 16.1% | 15 | 10.6% |  |  |
|  | 2,001 - 3,000 | 168 | 19.1% | 139 | 18.8% | 29 | 20.6% |  |  |
|  | 1,201 - 2,000 | 210 | 23.9% | 166 | 22.5% | 44 | 31.2% |  |  |
|  | 0 - 1,200 | 193 | 21.9% | 150 | 20.3% | 43 | 30.5% |  |  |
| Wealth Index | Richest | 179 | 20.3% | 175 | 23.7% | 4 | 2.8% | 44.5 | <0.001 |
|  | Richer | 173 | 19.7% | 153 | 20.7% | 20 | 14.2% |  |  |
|  | Middle | 180 | 20.5% | 144 | 19.5% | 36 | 25.5% |  |  |
|  | Poorer | 173 | 19.7% | 136 | 18.4% | 37 | 26.2% |  |  |
|  | Poorest | 175 | 19.9% | 131 | 17.7% | 44 | 31.2% |  |  |
| Job Loss | No | 392 | 44.5% | 347 | 47.0% | 45 | 31.9% | 10.2 | <0.001 |
|  | Yes | 488 | 55.5% | 392 | 53.0% | 96 | 68.1% |  |  |
| Food Price Increase | No | 356 | 40.5% | 326 | 44.1% | 30 | 21.3% | 25.6 | <0.001 |
|  | Yes | 524 | 59.5% | 413 | 55.9% | 111 | 78.7% |  |  |
| Pre-Covid Food Insecurity | No | 792 | 90.0% | 733 | 99.2% | 59 | 41.8% | 438.6 | <0.001 |
|  | Yes | 87 | 9.9% | 5 | 0.7% | 82 | 58.2% |  |  |

n= 880.
Data source: CHAMPS Ethiopia Lockdown Module data and CHAMPS Ethiopia Health and Demographic Surveillance data.
Notes: Nine observations were missing for head of household's education and occupation. One observation was missing for whether a household had experienced food insecurity prior to the onset of COVID-19. The "other" category for ethnicity included: Oromo, Somali, Gurage, Harari, Tigray, and "other". One observation was missing for whether a household had experienced food insecurity prior to the onset of COVID-19.

Since the onset of the pandemic, 15.7% of households reported not having enough food to eat, an increase of 5.8% from pre-pandemic levels; 59.5% of households reported an increase in food prices; and 55.5% of households reported having at least one family member lose a job during this period. Of the reasons reported by respondents for their household not being able to obtain sufficient food, roughly 88% reported that they couldn’t afford to buy more food, approximately 41% reported that they couldn’t get out to buy food, 23% of respondents reported fearing to go out and buy food, 20% of respondents reported that they couldn’t have groceries delivered, and 17% of respondents reported that stores didn’t have the food they wanted.

After adjusting for other characteristics (see Figure 2), households were more likely to report food insecurity if the household head identified as an ethnic group other than the Amhara (AOR = 3.28, 95% CI [1.08, 12.05]) or if their residence was located in an urban compared to a rural area (AOR = 2.62, 95% CI [1.10, 6.46]). Compared to households with 1 to 2 household members, larger households were more likely to have reported food insecurity; for example, for households with 7-8 members, the adjusted odds of reporting food insecurity were 5.42 times higher (95% CI [1.32, 23.35]). Compared to households with less than 1,200 Birr of monthly income, households with higher income were less likely to report food insecurity; for example, AOR = 0.18 (95% CI [0.05, 0.57]) for households with more than 4,600 Birr per month. Households in the wealthiest quintile were less likely to report food insecurity compared to the poorest households, AOR = 0.18, 95% CI [0.05, 0.57]). Households that observed increases in food prices were more likely to experience food insecurity (AOR = 2.20, 95% CI [1.11, 4.53]). Finally, after adjusting for other characteristics, households that reported food insecurity prior to the pandemic were more likely to report food insecurity during the pandemic (AOR = 423.03, 95% CI [139.44, 1685.07]). Interaction terms for evaluating effect modification between urban vs. rural residence and other characteristics were only statistically significant for job loss during the pandemic—urban households that had had a job loss were most likely to report insufficient food. All other associations were similar for urban and rural households.

**Figure 2.**
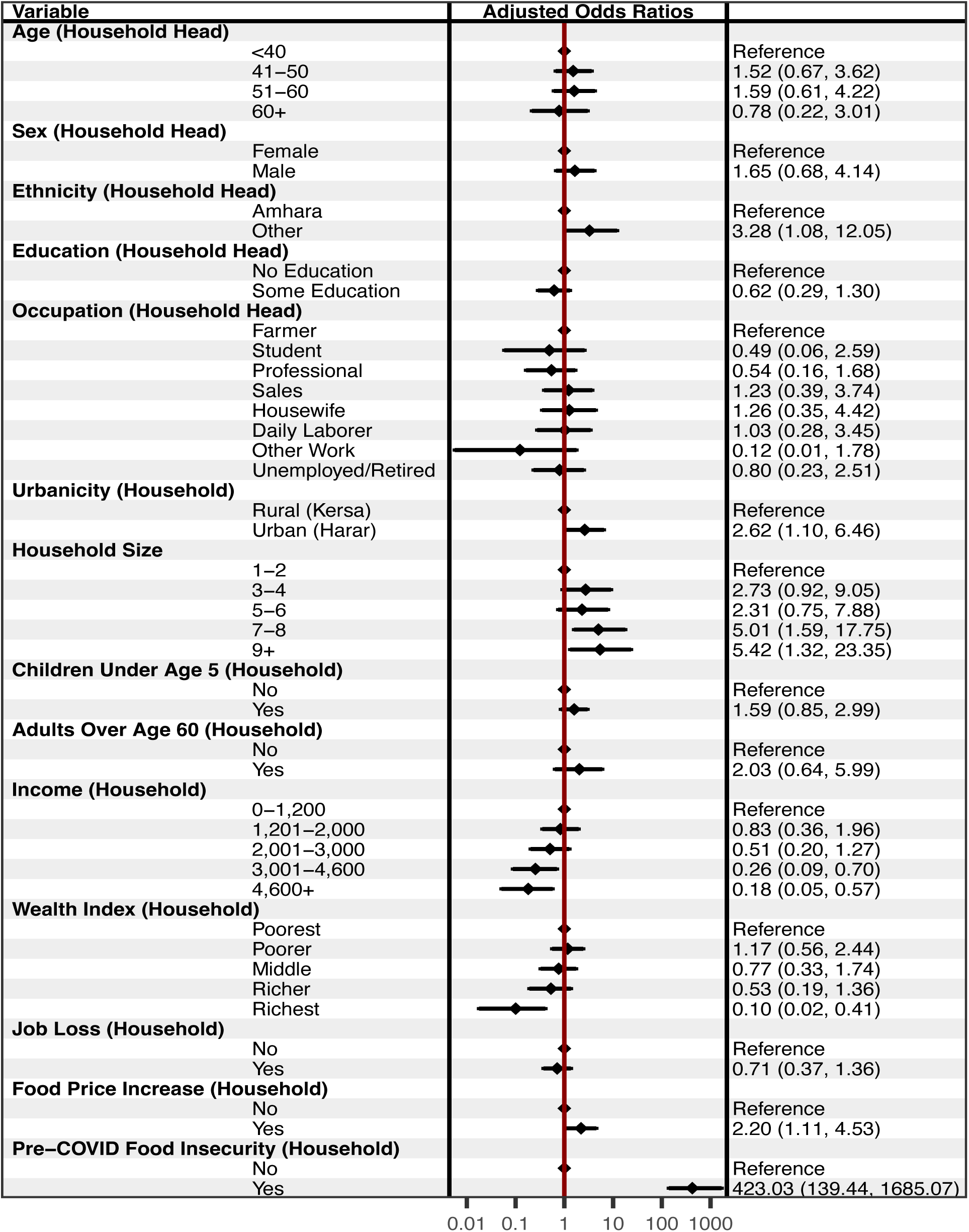
Adjusted Associations with Food Insecurity Among Households Living in Harar and Kersa, Ethiopia (n=870). The forest plot presents Adjusted Odds Ratios with 95% confidence intervals from an adjusted logistic regression model. Education and Occupation had 9 missing values.

In the secondary analysis that examined only those households that had indicated that they were food secure prior to the onset of the pandemic (see Figure 3), larger households were more likely to have reported food insecurity; for example, for households with 7-8 members, the adjusted odds of reporting food insecurity were 5.03 times higher (95% CI [1.47, 20.96]). Compared to households with less than 1,200 Birr of monthly income, households with higher income were less likely to report food insecurity; for example, AOR = 0.15 (95% CI [0.03, 0.56]) for households with more than 4,600 Birr per month. Richer households were less likely to report food insecurity compared to the poorest households, AOR = 0.16, 95% CI [0.05, 0.42]). Households that observed increases in food prices were more likely to experience food insecurity (AOR = 2.88, 95% CI [1.41, 6.27]). Ethnicity was dropped from the model due to lack of variation—only those households with a household head other than the Amhara became food insecure during the pandemic. Similarly, no households from the richest wealth quintile became food insecure during the pandemic.

**Figure 3.**
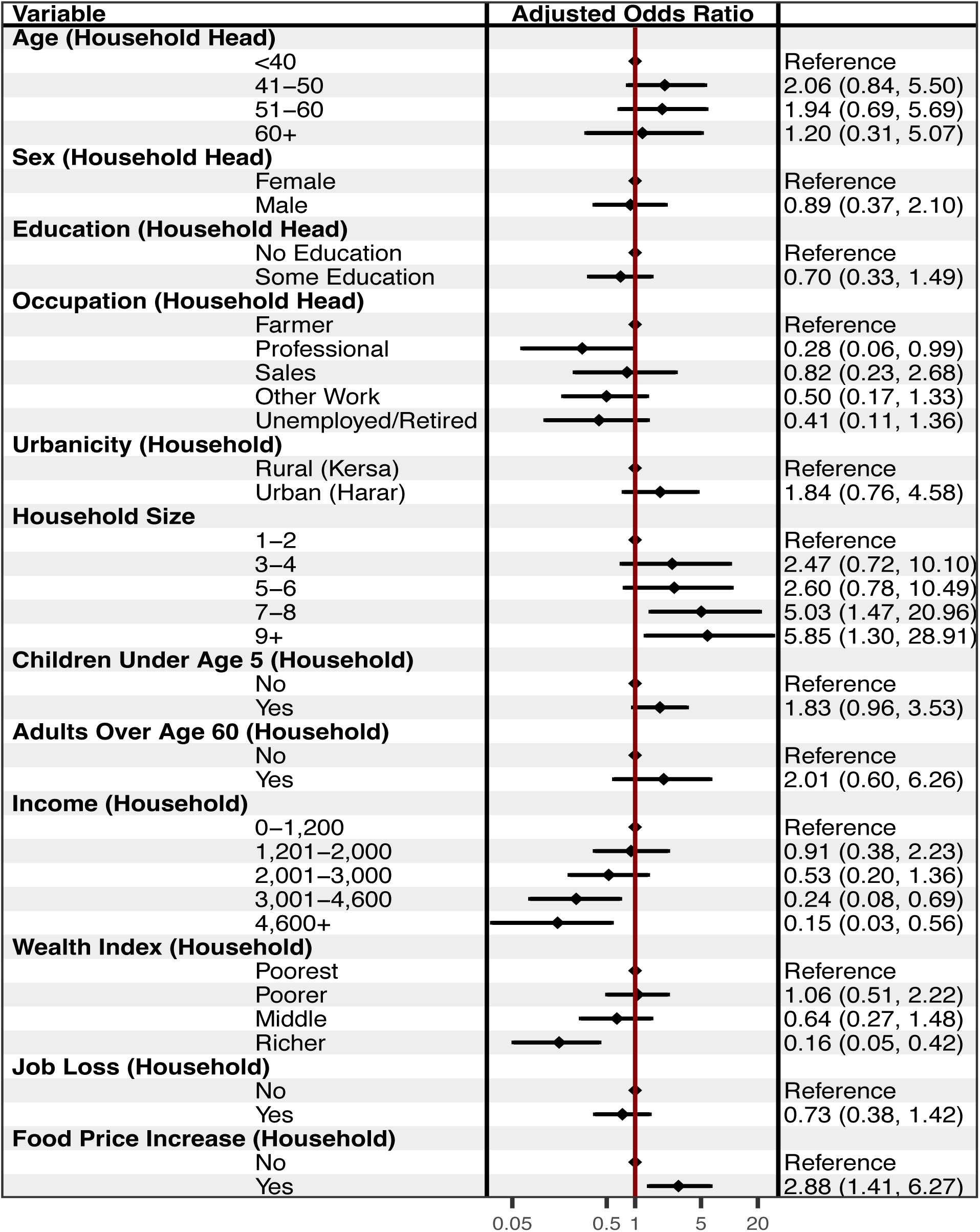
Adjusted Associations with Food Insecurity Among Previously Food Secure Households Living in Harar and Kersa, Ethiopia (n=870). The forest plot presents Adjusted Odds Ratios with 95% confidence intervals from an adjusted logistic regression model. Education and Occupation had 9 missing values.

## DISCUSSION

In this study we examined the overall prevalence and the development of household food insecurity during the pandemic in a rural and an urban community in eastern Ethiopia. Of the 10% of households that reported having difficulty obtaining sufficient food prior to the onset of the pandemic, 94% reported continued food insecurity during the pandemic. An additional 6% of households developed food insecurity during the pandemic (these households reported having difficulty obtaining sufficient food during the pandemic period, even though they did not have difficulties before). This level of increase is somewhat lower than national level increases reported in other studies, but consistent with findings that food insecurity increases were moderate in Oromia.[38] To contextualize this increase in food insecurity, we note that at the time these data were collected there were 39,779 households living within these communities. Applying the 6% increase to the entire population would correspond to an additional 2,387 households reporting food insecurity during the pandemic. The primary reason that households reported they were unable to obtain sufficient food was that it was too expensive. Additional reasons reported by a number of households included the inability to go out and buy food due to lockdown restrictions and fear to go out and buy food during the COVID-19 pandemic.

Overall, households were more likely to report experiencing food insecurity during the pandemic if they were already food insecure prior to COVID-19. The pandemic degenerated the food security of additional households that were vulnerable due to poverty or geospatial isolation; households were more likely to report experiencing food insecurity if they resided in an urban vs. a rural area, had a larger number of household members, had less monthly income, and were poorer compared to other households in the community. The pattern of association for both monthly income and household wealth is suggestive of a dose response to lower levels of household wealth and less monthly income. These associations are consistent with risk factors reported in studies from resource limited countries.[6, 11–13] Shocks during the pandemic associated with increased risk of food insecurity included increases in food prices and job loss; however, only increases in food prices were associated with households reporting that they were unable to obtain sufficient food after adjusting for pre-pandemic food insecurity.[6] Interventions should anticipate that households experiencing these shocks are at risk of food insecurity. Within the scope of the social vulnerability framework that we employed, we found both household level characteristics as well as community level characteristic associated with food insecurity; however, we found minimum between community variation for the associations between household level characteristics and food insecurity, the exception to these results is that the negative association between job loss and food insecurity was amplified in urban areas.

One limitation of this study, common to studies using HDSS data, is that the findings reported here are not generalizable outside of the communities under study.[14] As an observational study, other limitations include potential recall bias due to the extended length of time considered in the study or unmeasured variable bias; for example, having data on household participation in Ethiopia’s Productive Safety Net Program (PSNP) may have helped contextualize our results, particularly the relatively lower increase in food insecurity reported by households as other studies with access to this information have noted that increases in food insecurity during the pandemic were more moderate (less than 3%) for PSNP recipients.[38]

An additional limitation is that, given the broader goal of understanding how households’ circumstances changed during the pandemic, our measure of food insecurity is less comprehensive and does not adopt standard indices of food insecurity,[48] such as a household dietary diversity score.[49] Implementing a more comprehensive measure may have improved our ability to identify households that became food insecure during the pandemic in additional to the 6% that we have reported on in this study. As a cross-sectional study, we cannot draw causal inferences. Given findings from other studies that suggest hardships associated with COVID-19 lockdowns are temporary,[7] scholars should collect and analyze longitudinal data to evaluate changes in food insecurity over time. In the context of demographic surveillance systems, follow-up data collection using the same survey instrument is easy to attach to subsequent rounds of data collection already being fielded. A concern common to studies involving complex humanitarian crises is that we are unable to distinguish between the effects of the pandemic and possible impacts of the armed conflict in the northern regions of Ethiopia that was temporarily contiguous with the pandemic.[50] However, the political tension and civil conflict in Ethiopia have occurred primarily in the northernmost region of Tigray, which is more than 400 km from the HDSS.

By identifying factors associated with households’ inability to obtain sufficient food during the pandemic period, our findings have policy implications. Established food assistance programs are critical to support, even bolster, during pandemic periods as households previously identified as food insecure will continue to require assistance. However, our findings suggest that after taking other characteristics into consideration, households in urban areas were at higher risk for food insecurity. These findings point to the need for flexibility in applying food assistance programs that may have hitherto focused on assisting rural areas; intervention policies should consider the expansion of these programs to more urban areas in the context of pandemics that cause major shocks to food supply chains and other economic disruptions. Households in urban areas are particularly vulnerable to food insecurity when economic shocks result in loss of employment for household members. Given these findings, policies and practices that aim to mitigate the negative consequences of outbreaks should consider supplemental economic assistance to provide aid to households hit by broader economic shocks. These steps could mitigate the impact of lockdowns on more vulnerable households.

## Declarations

### Ethics approval and consent to participate

This study was conducted according to the guidelines in the Declaration of Helsinki; all procedures involving research study participants, including digital data collection using tablets that were programmed with the corresponding survey instruments, were approved by the Institutional Health Research Ethics Review Committee (IHRERC), College of Health and Medical Sciences, Harar Campus, Ethiopia; approval reference number Ref.No.IHRERC/127/2021. Written informed consent was obtained for participants who were able to read and write. For participants who were unable to read or write, the informed consent statement was read and oral informed consent from the participant was obtained, recorded, and witnessed. These procedures for obtaining written or oral informed consent were approved by the Institutional Health Research Ethics Review Committee (IHRERC), College of Health and Medical Sciences, Harar Campus, Ethiopia; approval reference number Ref.No.IHRERC/127/2021.

## Consent for publication

Not applicable.

## Availability of data and materials

Data from the survey questionnaire on impacts of the COVID-19 pandemic and related lockdowns, and supporting documentation, are accessible through the CHAMPS Population Surveillance data repository hosted by UNC Dataverse (https://doi.org/10.15139/S3/CZO1IX) [43]. The HDSS data analyzed in tandem with the COVID-19 survey data during the current study are available from Merga Dheresa of the corresponding author upon reasonable request.

## Authors’ contributions

J.M., S.C., N.A., M.D., and C.W. conceptualized and developed the methods for the study. M.D., N.A., T.G., G.M., and G.D. performed the data collection. J.M. and Z.M. carried out the formal analysis. J.M. wrote the original manuscript draft. J.M., M.D., Z.M., T.G., G.D., G.M., C.W., N.A., and S.C. reviewed and revised the manuscript. C.W., S.C., and N.A provided supervision for the study. C.W. acquired funding.

## Data Availability

All data produced in the present study are available upon reasonable request to the authors

## Acknowledgements

Not applicable.

## Disclaimers

The findings and conclusions in this report are those of the authors and do not necessarily represent the views of the US Centers for Disease Control and Prevention.

## SUPPLEMENT A. WEALTH INDEX GENERATION

A wealth index was generated based on a collection of assets and construction materials for the main dwelling of a given household. To generate the index, we followed recommendations from the DHS and the World Food Programme (WFP) that summarize steps for calculating an asset based wealth index,[51, 52] including coding instructions for Stata,[53] and an adaptation of this coding process implemented in R.[31, 50] Using these documents to guide us, we generated our wealth index by identifying a list of household assets for inclusion in our index computation, recoded all household assets into dichotomous variables; recoded dwelling materials into improved vs. non-improved dichotomous variables; divided our sample into rural and urban subsamples and assessed level of representation of a given variable within the rural and urban subsamples (per WFP recommendations, a given variable was included in further calculations if percent ownership ranged between 5 and 95 percent); employed principal components analysis with varimax rotation to calculate component scores for those households living in either rural or urban areas, which explained 45% of the variation in both subsamples; extracted and combined the PCA scores of the first component from the urban and rural subsamples; and finally organized the scores into wealth quintiles to generate a composite asset index. The resulting wealth quintiles are presented in Figure (4). The distribution of assets owned by households as well as the materials used for constructing a household’s residence are presented as percentages in Figure (5).

**Figure 4.**
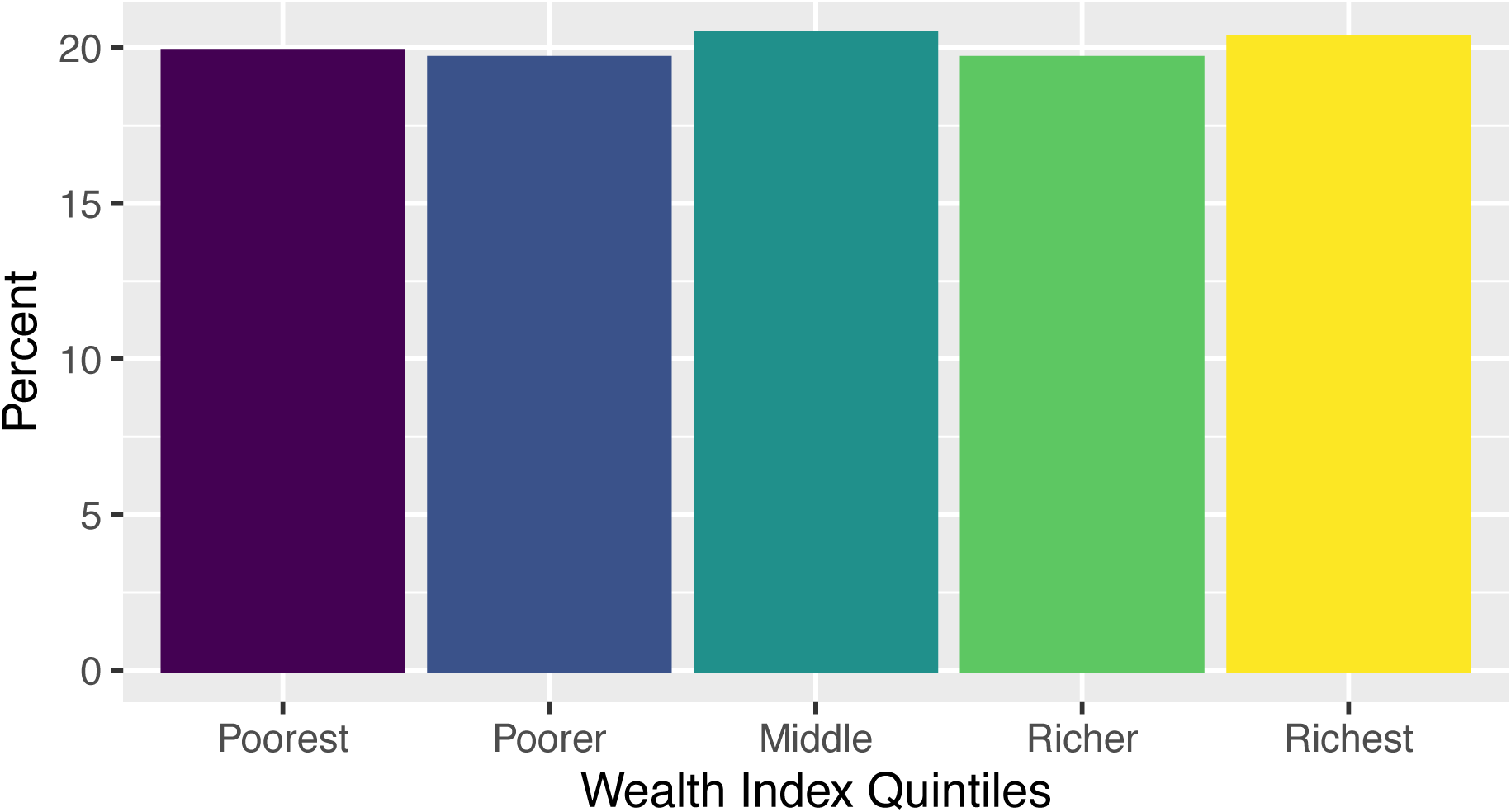
Household Wealth Index Distribution

**Figure 5.**
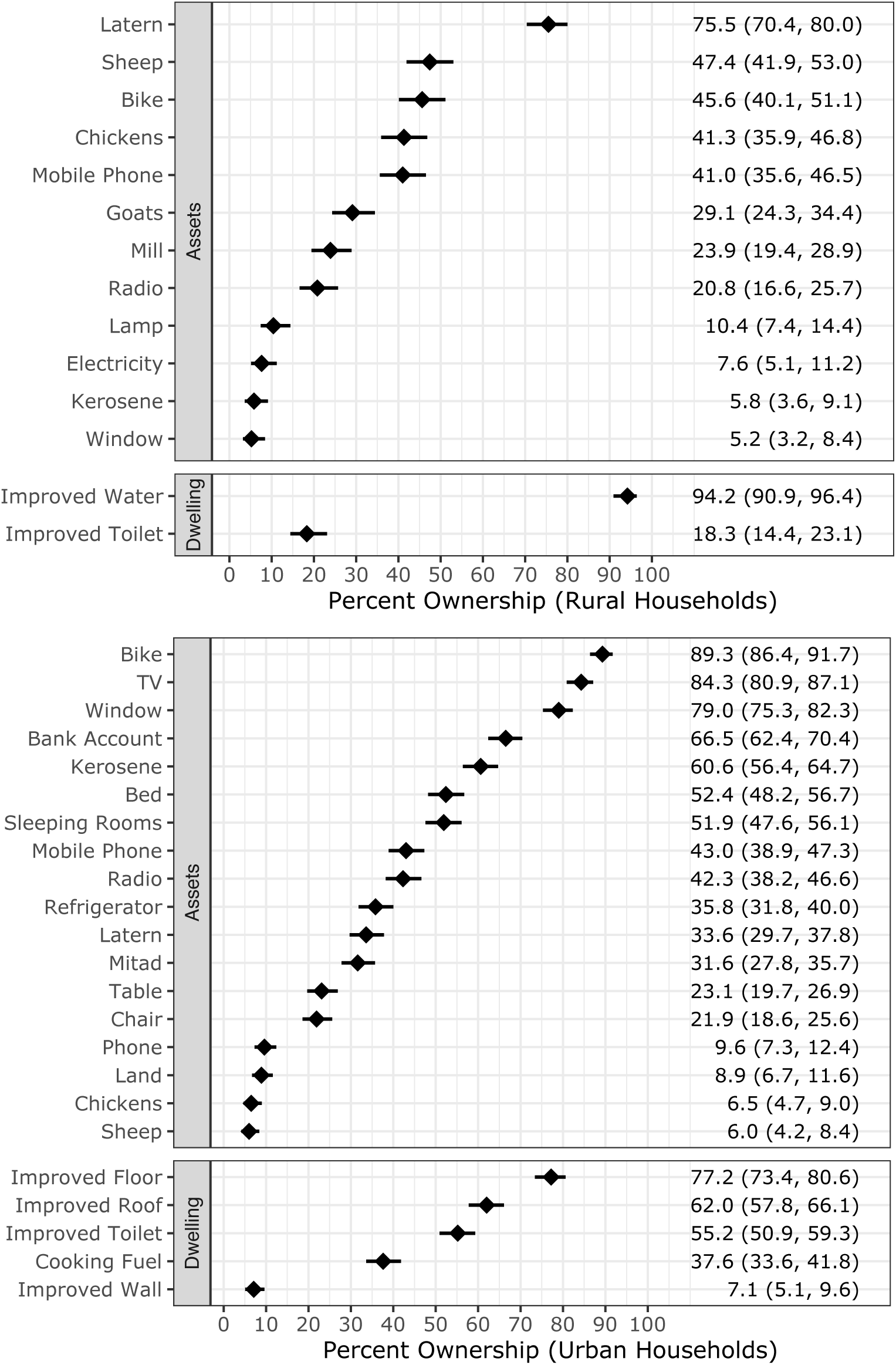
Households Asset Ownership and Dwelling Construction Materials included in Wealth Index

## Footnotes

1 Based on exchange rates at the time of the survey, this is roughly equivalent to 1 = 88+ USD (reference), 2 = 55 to 87 USD, 3 = 30 to 54 USD, and 4 = Less than 30 USD.

2 For information on the principal components analysis used to compute the index, see Supplemental Materials.

3 Based on responses to the question: “Before mid-March 2020, did it ever happen that your household did not have enough food to eat?” from the CHAMPS Ethiopia Lockdown Module.

## REFERENCES

1. Wang H, Paulson KR, Pease SA, Watson S, Comfort H, Zheng P, Aravkin AY, Bisignano C, Barber RM, Alam T: Estimating excess mortality due to the COVID-19 pandemic: a systematic analysis of COVID-19-related mortality, 2020–21. The Lancet 2022.

2. Baldwin RE, Weder B: Mitigating the COVID Economic Crisis: Act Fast and Do Whatever It Takes: CEPR press; 2020.

3. Picchioni F, Goulao LF, Roberfroid D: The Impact of COVID-19 on Diet Quality, Food Security and Nutrition in Low and Middle Income Countries: A Systematic Review of the Evidence. Clinical Nutrition 2021.

4. Osendarp S, Akuoku JK, Black RE, Headey D, Ruel M, Scott N, Shekar M, Walker N, Flory A, Haddad L: The COVID-19 Crisis Will Exacerbate Maternal and Child Undernutrition and Child Mortality in Low-and Middle-Income Countries. Nature Food 2021, 2(7):476–484.

5. Akseer N, Kandru G, Keats EC, Bhutta ZA: COVID-19 Pandemic and Mitigation Strategies: Implications for Maternal and Child Health and Nutrition. The American Journal of Clinical Nutrition 2020, 112(2):251–256.

6. Onyango EO, Crush J, Owuor S: Preparing for COVID-19: Household Food Insecurity and Vulnerability to Shocks in Nairobi, Kenya. PLoS ONE 2021, 16(11):e0259139.

7. Hirvonen K, de Brauw A, Abate GT: Food Consumption and Food Security During the COVID-19 Pandemic in Addis Ababa. American Journal of Agricultural Economics 2021, 103(3):772–789.

8. Ayanlade A, Radeny M: COVID-19 and Food Security in Sub-Saharan Africa: implications of Lockdown during Agricultural Planting Seasons. npj Science of Food 2020, 4(1):1–6.

9. Hirvonen K, Abate GT, de Brauw A: Survey Suggests Rising Risk of Food and Nutrition Insecurity in Addis Ababa, Ethiopia, as COVID-19 Restrictions Continue. IFPRI book chapters 2020:46–49.

10. Chitiga M, Henseler M, Mabugu RE, Maisonnave H: How COVID-19 pandemic worsens the economic situation of women in South Africa. The European Journal of Development Research 2021:1–18.

11. Fikire AH, Zegeye MB: Determinants of Rural Household Food Security Status in North Shewa Zone, Amhara Region, Ethiopia. The Scientific World Journal 2022, 2022.

12. Manfrinato CV, Marino A, Condé VF, Maria do Carmo PF, Stedefeldt E, Tomita LY: High prevalence of food insecurity, the adverse impact of COVID-19 in Brazilian favela. Public health nutrition 2021, 24(6):1210–1215.

13. Banna M, Al H, Sayeed A, Kundu S, Kagstrom A, Sultana M, Begum MR, Khan MSI: Factors associated with household food insecurity and dietary diversity among day laborers amid the COVID-19 pandemic in Bangladesh. BMC nutrition 2022, 8(1):1–11.

14. Clark S, Wakefield J, McCormick T, Ross M: Hyak Mortality Monitoring System: Innovative Sampling and Estimation Methods–Proof of Concept by Simulation. Global Health, Epidemiology and Genomics 2018, 3.

15. Cutter SL, Boruff BJ, Shirley WL: Social vulnerability to environmental hazards. In: Hazards vulnerability and environmental justice. edn.: Routledge; 2012: 143–160.

16. Cutter SL, Barnes L, Berry M, Burton C, Evans E, Tate E, Webb J: A place-based model for understanding community resilience to natural disasters. Global environmental change 2008, 18(4):598–606.

17. Summit W: Rome declaration on world food security and world food summit. Plan of action Roma, FAO 1996, 43.

18. Elavarasan RM, Pugazhendhi R: Restructured Society and Environment: A Review on Potential Technological Strategies to Control the COVID-19 Pandemic. Science of The Total Environment 2020, 725:138858.

19. Roberton T, Carter ED, Chou VB, Stegmuller AR, Jackson BD, Tam Y, Sawadogo-Lewis T, Walker N: Early Estimates of the Indirect Effects of the COVID-19 Pandemic on Maternal and Child Mortality in Low-Income and Middle-Income Countries: A Modeling Study. The Lancet Global Health 2020, 8(7):e901–e908.

20. Dowd JB, Andriano L, Brazel DM, Rotondi V, Block P, Ding X, Liu Y, Mills MC: Demographic Science Aids in Understanding the Spread and Fatality Rates of COVID-19. Proceedings of the National Academy of Sciences 2020, 117(18):9696–9698.

21. Onder G, Rezza G, Brusaferro S: Case-Fatality Rate and Characteristics of Patients Dying in Relation to COVID-19 in Italy. Jama 2020, 323(18):1775–1776.

22. Ruan Q, Yang K, Wang W, Jiang L, Song J: Clinical Predictors of Mortality Due to COVID-19 Based on an Analysis of Data of 150 Patients From Wuhan, China. Intensive care medicine 2020, 46(5):846–848.

23. Goshu D, Ferede T, Diriba G, Ketema M: Economic and Welfare Effects of COVID-19 and Responses in Ethiopia: Initial Insights. In.: Ethiopian Economic Policy Research Institute; 2020.

24. Shigute Z, Mebratie AD, Alemu G, Bedi A: Containing the Spread of COVID-19 in Ethiopia. Journal of Global Health 2020, 10(1).

25. Baye K: COVID-19 Prevention Measures in Ethiopia: Current Realities and Prospects, vol. 141: International Food Policy Research Institute; 2020.

26. Spielman SE, Tuccillo J, Folch DC, Schweikert A, Davies R, Wood N, Tate E: Evaluating Social Vulnerability Indicators: Criteria and Their Application to the Social Vulnerability Index. Natural Hazards 2020, 100(1):417–436.

27. Muir JA, Cope MR, Angeningsih LR, Jackson JE, Brown RB: Migration and Mental Health in the Aftermath of Disaster: Evidence From MT. Merapi, Indonesia. International journal of environmental research and public health 2019, 16(15):2726.

28. Muir JA, Cope MR, Angeningsih LR, Jackson JE: To Move Home or Move On? Investigating the Impact of Recovery Aid on Migration Status as a Potential Tool for Disaster Risk Reduction in the Aftermath of Volcanic Eruptions in Merapi, Indonesia. International Journal of Disaster Risk Reduction 2020, 46:101478.

29. Perry RW: What is a Disaster? In: Handbook of Disaster Research. edn.: Springer; 2007: 1–15.

30. Flanagan BE, Gregory EW, Hallisey EJ, Heitgerd JL, Lewis B: A Social Vulnerability Index for Disaster Management. Journal of Homeland Security and Emergency Management 2011, 8(1).

31. Muir JA: Another mHealth? Examining Motorcycles as a Distance Demolishing Determinant of Health Care Access in South and Southeast Asia. Journal of Transport & Health 2018, 11:153–166.

32. Sanders SR, Muir JA, Brown RB: Overcoming Geographic Penalties of Inequality: The Effects of Distance-Demolishing Technologies on Household Well-Being in Vietnam. Asian Journal of Social Science 2018, 46(3):260–280.

33. Muir JA, Deresa M, Madewell ZJ, Getachew T, Mangesha G, Whitney CG, Assefa N, Cunningham SA: Household Hardships during the COVID-19 Pandemic: Examining Household Vulnerability and Responses to Pandemic Related Shocks in Eastern Ethiopia. medRxiv 2023:2023.2002. 2001.23285322.

34. Bedeke SB: Food insecurity and copping strategies: a perspective from Kersa district, East Hararghe Ethiopia. Food Science and Quality Management 2012, 5(3):19–27.

35. Gebru M, Remans R, Brouwer ID, Baye K, Melesse MB, Covic N, Habtamu F, Abay AH, Hailu T, Hirvonen K: Food systems for healthier diets in Ethiopia: Toward a research agenda. IFPRI Discussion Paper 2018.

36. Haile D, Seyoum A, Azmeraw A: Food and nutrition security impacts of resilience capacity: Evidence from rural Ethiopia. Journal of Agriculture and Food Research 2022, 8:100305.

37. Clay DC, Molla D, Habtewold D: Food aid targeting in Ethiopia: A study of who needs it and who gets it. Food policy 1999, 24(4):391–409.

38. Abay K, Berhane G, Hoddinott J, Tafere K: COVID-19 and food security in Ethiopia: do social protection programs protect? 2020.

39. Juntunen L: Addressing Social Vulnerability to Hazards. University of Oregon Eugene; 2004.

40. Assefa N, Oljira L, Baraki N, Demena M, Zelalem D, Ashenafi W, Dedefo M: HDSS profile: the Kersa Health and Demographic Surveillance System. International Journal of Epidemiology 2016, 45(1):94–101.

41. Cunningham SA, Shaikh NI, Nhacolo A, Raghunathan PL, Kotloff K, Naser AM, Mengesha MM, Adedini SA, Misore T, Onuwchekwa UU: Health and Demographic Surveillance Systems within the Child Health and Mortality Prevention Surveillance Network. Clinical Infectious Diseases 2019, 69(Supplement_4):S274–S279.

42. Nhacolo A, Madewell ZJ, Muir JA, Sacoor C, Xerinda EG, Matsena T, Bassat Q, Whitney CG, Mandomando I, Cunningham SA: Knowledge of COVID-19 Symptoms, Transmission, and Prevention: Evidence from Health and Demographic Surveillance in Southern Mozambique. medRxiv 2023:2023.2003. 2031.23288026.

43. Dheresa M, Muir JA, Madewell ZJ, Getachew T, Daraje G, Mengesha G, Whitney CG, Assega N, Cunningham SA: COVID-19 Impact Data for the CHAMPS HDSS Network: Data from Harar and Kersa, Ethiopia. In., V1 edn: UNC Dataverse; 2023.

44. Cunningham SA, Muir JA: Data Cleaning. In: The Cambridge Handbook of Research Methods and Statistics for the Social and Behavioral Sciences: Volume 1: Building a Program of Research (Cambridge Handbooks in Psychology, pp 443-467). Volume 1, edn. Edited by Nichols AL, Edlund JE. Cambridge: Cambridge University Press; 2023.

45. R Core Team: R: A Language and Environment for Statistical Computing. In. Vienna, Austria: R Foundation for Statistical Computing; 2022.

46. Yeh J, D’Amico F: Forest plots: data summaries at a glance. Journal of Family Practice 2004, 53(12):1007–1008.

47. Lewis SC, Keerie C, Assi V: Forest Plot. In: Wiley StatsRef: Statistics Reference Online. edn.: 1–6.

48. Maxwell D, Vaitla B, Coates J: How do indicators of household food insecurity measure up? An empirical comparison from Ethiopia. Food policy 2014, 47:107–116.

49. Swindale A, Bilinsky P: Household dietary diversity score (HDDS) for measurement of household food access: indicator guide. *Washington, DC*: Food and Nutrition Technical Assistance Project, Academy for Educational Development 2006.

50. Muir JA: Societal Shocks as Social Determinants of Health: The Ohio State University; 2021.

51. Rutstein SO: Steps to Constructing the New DHS Wealth Index. *Rockville, MD*: ICF International 2015.

52. Hjelm L, Mathiassen A, Miller D, Wadhwa A: Creation of a wealth index. United Nations World Food Programme 2017.

53. Fry K, Firestone R, Chakraborty NM: Measuring Equity With Nationally Representative Wealth Quintiles. Washington DC 2014.

